# Protocol for an observational study evaluating new approaches to modelling diagnostic information from large administrative hospital datasets

**DOI:** 10.1101/19011338

**Authors:** Thomas E Cowling, David A Cromwell, Linda D Sharples, Jan van der Meulen

## Abstract

**Introduction:** The Charlson and Elixhauser indices define sets of conditions used to adjust for patients’ comorbidities in administrative hospital data. A strength of these indices is the parsimony that results from including only 19 and 30 conditions respectively, but the conditions included may not be the ones most relevant to a specific outcome and population. Our objectives are to: (1) test an approach to developing parsimonious indices for the specific outcome and populations being studied, while comparing performance to the Charlson and Elixhauser indices; and (2) evaluate several approaches that involve models with more diagnosis-related terms and aim to improve prediction performance by capturing more of the information in large datasets.

**Methods and analysis:** This is a modelling study using a linked national dataset of administrative hospital records and death records. The study populations are patients admitted to hospital for acute myocardial infarction, hip fracture, or major surgery for colorectal cancer in England between 1 January 2015 and 31 December 2017. The outcome is death within 365 days of the date of admission (acute myocardial infarction and hip fracture) or procedure (colorectal surgery). In the ‘First analysis’, prognostic indices will be developed based on the presence/absence of individual ICD-10 codes in patients’ medical histories. Logistic regression will be used to estimate associations with a full set of sociodemographic and diagnostic predictors from which reduced models (with fewer diagnostic predictors) will be produced using a step-down approach. In the ‘Second analysis’, models will also account for the timing that each ICD-10 code was last recorded and allow for non-linear relationships and interactions between conditions and the timings of records. Validation will include an overall measure of performance (scaled Brier score) and measures of discrimination (*c*-statistic) and calibration (such as the Integrated Calibration Index) in bootstrap or cross-validation samples. Sensitivity analyses will include varying the length of medical history analysed, using a comparator that combines the Charlson and Elixhauser sets of conditions, and aggregating ICD-10 codes into clinical groups.

## Introduction

In 1987, Charlson *et al*.^1^ proposed a set of comorbidities that could be used to predict patient mortality in longitudinal studies. The corresponding article is now the 3^rd^ most cited article of all time in the ‘Medicine, General and Internal’ category of the Web of Science^2^ with almost 30,000 citations (Google Scholar). The wider set of comorbidities proposed in 1998 by Elixhauser *et al*.^3^, for use with large administrative hospital datasets, has almost 6,000 citations. These two indices are a cornerstone of modern clinical research and healthcare evaluation: they are often used to assess and reduce confounding of outcomes between trial arms,^4-6^ observed treatment groups,^7-9^ and healthcare providers.^10-12^ They are also often included in prediction models.^13-15^

These indices are likely to be popular partly because they are highly transparent, reproducible, and parsimonious. The Charlson^1^ and Elixhauser^3^ indices include 19 and 30 conditions, respectively, which have well-established International Classification of Diseases (ICD) coding methods.^16^ The weighted number of conditions is often used as a summary measure.^1,17^

However, the relatively small number of conditions included in each index may not account for the ones most relevant to a specific outcome and population,^18^ thus limiting the degree to which confounding is reduced. Several authors have therefore suggested approaches that include more conditions.^19-22^ For example, in 2005, Holman *et al*.^23^ used the 100 commonest ICD codes in the population to define candidate predictor variables. While this approach performed better than the Charlson index in its development study,^23^ it has not been widely used (with less than 100 citations on Google Scholar). This may be partly because the large number of variables is likely to increase the risk that models fitted in smaller datasets do not generalise well to other datasets. It is also more time-consuming to apply and present a larger model in different studies.

This highlights the need for a new approach to developing prognostic indices that considers a large number of candidate conditions but produces a final model with a relatively small number of conditions that account for most of the explainable variation in the outcome. This model is adapted to the specific outcome and population being studied and should share the strengths of the Charlson and Elixhauser indices given above, especially their parsimony.

Several studies focusing on risk prediction in large datasets, rather than a comorbidity index in itself, have developed particularly large models with many more diagnosis-related predictors. These models have accounted for the timing of diagnosis records^24-26^ and were estimated using machine learning methods that do not assume a simple functional form for associations between outcomes and predictors.^24-28^ This could help these approaches to better predict outcomes than more conventional methods, but this has not been tested or quantified empirically in the existing literature. Moreover, more complex models may be less interpretable and reproducible than simpler ones, which may negate any benefits in terms of prediction performance.

In this study, we will: (1) test an approach to developing parsimonious prognostic indices, based on ICD-10 codes, for the specific outcome and populations being studied, while comparing performance to the Charlson and Elixhauser indices; and (2) evaluate several approaches that involve models with more diagnosis-related terms and aim to improve prediction performance by capturing more of the information in large datasets.

The study outcome and populations will be 1-year mortality of acute myocardial infarction, hip fracture, and major colorectal cancer surgery patients in England. We have chosen to analyse specific groups rather than all inpatients as most studies using comorbidity methods focus on clinical populations defined by disease, events, or interventions, so comparisons of methods are most informative (and most common^18^) when done within clinical groups.^23^

We chose these three groups to illustrate the general applicability of the approaches. The groups correspond to a large number of admissions per year but vary in terms of the clinical areas affected, the conditions that coexist, and mortality rates. Most deaths amongst patients admitted for hip fracture are due to other conditions, both in the short and longer terms,^29,30^ with a 1-year mortality rate of approximately 30%.^31^ In contrast, acute myocardial infarction presents a greater risk of death in itself; most deaths within 30 days of admission for this condition are due to the primary event.^32^ Unlike these two conditions, most major surgery for colorectal cancer is planned and the 1-year mortality rate is less than 10%.^36^ The approaches will therefore be tested in a heterogeneous group of conditions where performance is more likely to vary.

## Methods

### Study populations

We will conduct a modelling study using a large national dataset of administrative hospital records. The Hospital Episode Statistics Admitted Patient Care dataset includes all inpatient hospital care funded by the National Health Service (NHS) in England.^37^

The three study populations are patients admitted to hospital for acute myocardial infarction (ICD-10: I21-22^38,39^), hip fracture (S72.0-S72.2^40,41^), or major surgery for colorectal cancer (C18-20; OPCS-4: H04-11, H29, H33, X14^42-45^). Acute myocardial infarction and hip fracture patients will be identified from the ICD-10 code recorded as the primary diagnosis for the first episode in each admission. Colorectal surgery patients will be identified from the primary diagnosis of any episode with a relevant OPCS-4 code recorded for the main procedure of that same episode.

We will analyse patients aged 18 years or older in the acute myocardial infarction and major colorectal surgery populations and aged 60 years or older^41^ in the hip fracture population. We will include those with a relevant admission from 1 January 2015 to 31 December 2017.

Each record in Hospital Episode Statistics corresponds to an ‘episode’, defined as a continuous period of hospital care under the same consultant doctor. Each episode has 20 diagnosis fields which contain ICD-10 codes corresponding to diagnoses recorded for that specific episode. The first field contains the primary diagnosis—the main condition treated or investigated during that episode. The other 19 fields contain secondary diagnoses. Three-character categories of ICD-10 codes define single conditions or groups of diseases with a common characteristic; fourth characters are used variably to define sites, subtypes, and causes.^46^

### Outcome

The outcome variable is death within 365 days of the date of admission (acute myocardial infarction and hip fracture) or procedure (major colorectal surgery). Death in hospital or within 30 days and 365 days of admission are the commonest outcome variables in studies comparing comorbidity methods^18,47^; we will focus on the 365 days period (as in Charlson *et al*.^1^) as in-hospital and 30-day mortality may be much more strongly affected by the primary event than other conditions. In addition, 30-day mortality after colorectal excision is less than 3%.^36^ If a patient has more than one index admission of the same type (such as two admissions for hip fracture), the earliest date of these admissions from 2015 to 2017 will be taken as the index date.

We will identify dates of death from Civil Registrations Data.^48^ We have these records up to 31 December 2018, providing complete follow-up for the outcome. Death records were linked to Hospital Episode Statistics using deterministic linkage on each patient’s unique NHS number, date of birth, sex, and postcode.

### First analysis – Parsimonious models

#### Definition of predictors

We will analyse diagnosis codes from episodes that started within 365 days before the index date of each patient. We will use the three-character categories of ICD-10 codes only (excluding fourth characters) to increase the frequencies of predictors so that model predictions are more reliable. In each population, we will exclude code categories recorded for less than 0.5% of patients as these codes are so rare that they are unlikely to explain much variation in outcomes.^25,49^

We will define a binary predictor variable for each code that denotes whether that code was recorded or not within 365 days before a patient’s index date. We chose this 1-year ‘look-back period’ rather than only using information from the index admission as the former approach can also be applied in non-inpatient populations and some studies suggest that it improves model performance.^18^

Patient age (coded as a continuous variable) at the index date, sex, and socioeconomic status will be predictors in all models, as is common when comparing comorbidity indices.^18,47^ Socioeconomic status will be measured by the national rank of the Index of Multiple Deprivation score for the lower super output area in which a patient lived; these areas are ranked from 1 (most deprived) to 32,482 (least deprived) and have population sizes between 1,000 and 3,000.^50^

#### Model estimation

We will first estimate associations between the outcome and the full set of predictors as the maximum likelihood estimates from logistic regression. Interaction terms will not be considered. We will then develop a reduced model that approximates this full model using the backwards step-down approach proposed by Harrell.^51,52^

First in this approach, the predicted log-odds of the outcome for each patient is calculated from the coefficients of the full model. Second, an ordinary least squares regression model is fitted between these values and the full set of predictors (with an *R*^2^ value of 1 by definition). Third, *R*^2^ is calculated for a series of models in which each successive model has an additional predictor removed, in a backwards step-down process based on likelihood ratio estimates.^51,53^ This process will stop when all predictors have been removed. Fourth, the model with the fewest predictors and an *R*^2^ value equal to or greater than 0.95 will be selected as the final reduced model. This model will explain at least 95% of the variation in the outcome explained by the full set of predictors.

#### Comparators

We will compare this resulting prediction approach to the Charlson^1^ and Elixhauser^3^ methods, as systematic reviews found them to be the most commonly used comorbidity indices by far.^18,47^

An established list of ICD-10 codes^16^ will be used to identify whether each condition included in these indices was recorded or not within 365 days before the index date. This list categorises the Charlson^1^ and Elixhauser^3^ conditions into 17 and 31 groups, respectively; 14 of the 17 Charlson groups are also included in the Elixhauser groups.^54^ We will estimate associations between the outcome and each condition using multivariable logistic regression, separately for the two sets of conditions. We will model a binary predictor variable for each condition rather than an overall weighted summary measure, as optimal weights differ across datasets, populations, outcomes, and time.^3,33^ Systematic reviews of comorbidity indices suggest that modelling conditions using this approach generally improves prediction performance.^18,47^

### Second analysis – Larger models

#### Use of diagnosis timings as predictors

The analysis described above will model the presence or absence of each ICD-10 code within a year before the index dates. In the second analysis, we will use a longer look-back period of five years and define an additional predictor for each ICD-10 code that records the number of days before the index date that the code was last recorded (from 0 to 1825 days). Logistic regression will be used to estimate the coefficients of the following model:

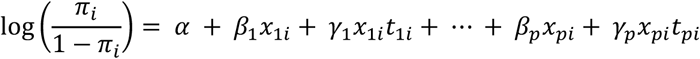

where *π*_*i*_ is the predicted probability of death for individual *i*, each of the variables *x*_*ki*_ represents a binary indicator denoting the presence (1) or absence (0) of the *k*th ICD-10 code (in the look-back period) for individual *i* and *t*_*ki*_ is a continuous variable denoting the corresponding number of days between the last record of that code and the date of the index admission/procedure. The model parameters are interpreted as follows:

- *α* represents the log-odds of death within 365 days for a person without any ICD-10 codes recorded
- *β*_*k*_, *k* = 1, …, *p* is the change in log-odds of death for the *k*th ICD-10 code on the day of the index admission/procedure
- *γ*_*k*_, *k* = 1, …, *p* is the change in log-odds of death per day for the interval between the last record of a given ICD-10 code and the index admission/procedure

Patient age, sex, and socioeconomic status will also be included as predictors (as before) and are omitted from the equation above only for clarity.

This equation assumes linear relationships between the timings that ICD-10 codes were last recorded and the log-odds of death. To relax this assumption, we will also fit generalised additive models that include non-linear associations for the predictors using smoothing splines.^55,56^

#### Use of boosted trees to account for interactions

A potential limitation of the approaches described above for the first and second analyses is that they do not model interactions between conditions. As conditions could modify each other’s effects on prognosis, we will also test the performance of boosted tree models. These models have the potential benefit of modelling interactions between conditions without these interactions having to be pre-specified, unlike when using conventional logistic regression for example. The approach involves fitting a series of decision trees to the data sequentially with each tree attempting to improve on the predictions of the previous tree (termed ‘boosting’).

We will use the gradient boosted tree approach of Friedman *et al*.^57-59^ as implemented in the XGBoost^60^ R package (explained accessibly elsewhere^61-63^). This is a popular statistical learning algorithm that has outperformed other methods in large routine healthcare datasets with many predictors.^24,26,28^ The tuning parameters of the algorithm are given in Appendix 1.

### Validation methods

For all analyses, Brier scores will be calculated to assess the overall predictive performance of the approaches.^64^ These scores equal the mean of squared differences between the predicted probability of death and the observed death outcome. Since Brier scores are affected by the overall risk of the outcome, we will scale these scores (Brierscaled) to account for the maximum possible value (Briermax) given the overall risk of death: Brierscaled = 1 – Brier/Briermax.^65^ Larger values of Brierscaled are better: this measure equals 100% when predictions are perfect and 0% when the predicted probability of death for each patient equals the overall risk of death in the population.

To assess discrimination, we will calculate the *c-*statistic.^66^ This measure equals the probability that a randomly chosen patient who died had a greater predicted probability of death than a randomly chosen patient who did not die; it assesses the rank order of predictions. A perfect model would have a value of 1 and a set of random predictions would have a value of 0.5.

To assess model calibration, we will calculate calibration-in-the-large, calibration slopes,^67^ and the Integrated Calibration Index (ICI).^68^ Calibration-in-the-large is the difference in means between predicted probabilities and observed death outcomes, such that perfect models have values of zero. Calibration slopes equal the coefficient of the predicted log-odds of death in a logistic regression model with the log-odds of death as the outcome variable and no other predictors; perfect models have slopes equal to one. The ICI is a weighted average of absolute differences between a calibration curve and the diagonal line of perfect calibration^68^; the perfect ICI is zero.

In the ‘First analysis’, we will calculate the above measures in the entire dataset used to fit the regression models (‘apparent performance’) and also optimism-corrected estimates of these measures using the bootstrapping procedure given in the TRIPOD guidelines^69-71^; 500 bootstrap samples will be used with all modelling steps repeated in each sample.

In the ‘Second analysis’, the models will take longer to estimate due to the larger number of parameters and bootstrapping will be too time-consuming, so a different validation process will be used. In each population, we will randomly allocate 75% of patients to a ‘training’ dataset and the other 25% to a ‘test’ dataset. We will first calculate the above performance measures in the training set using five repeats of 10-fold cross-validation. The results will be used to select the tuning parameters of the boosted trees that minimise the negative log-likelihood. We will then fit the final models on the whole training dataset and evaluate their prediction performance in the test dataset.

### Sensitivity analyses

Five analyses will test the sensitivity of results to changes in the methods described above for the ‘First analysis’. First, ICD-10 code categories recorded for less than 1% of patients (rather than 0.5%) will be excluded. Second, we will examine the performance of the full models with diagnostic predictors based on the index episode only or a 3-year look-back period (instead of a 1-year period). Third, we will fit a model using the conditions included in either of the Charlson and Elixhauser indices,^19,22^ using the most inclusive ICD-10 code definition of conditions in both indices.^54^ Fourth, we will aggregate ICD-10 codes into the groups of the Clinical Classification Software (CCS) scheme,^72,73^ excluding groups recorded for less than 0.5% of patients, and re-estimating the full models. Fifth, we will assess changes in coefficients of the full models when penalised maximum likelihood estimation^51,74^ is used; the penalty factor will be chosen to maximise a modified Akaike’s Information Criterion (AIC).^51,71,74^

The results of these analyses will inform which sensitivity analyses, if any, to also conduct for the approaches given under ‘Second analysis’ above.

We will follow the ‘Transparent Reporting of a multivariable prediction model for Individual Prognosis or Diagnosis’ (TRIPOD) guidelines for reporting the study.^70^ The corresponding articles will provide code for estimating the different models in R. Data management and preparation will be done using Stata (v15)^75^ and R (v3.5)^76^ will be used for all statistical analysis.

### Implications for research

This study will evaluate new approaches to modelling diagnostic information from large administrative hospital datasets, using mortality as the outcome. These approaches could be particularly useful for risk adjustment when comparing patient outcomes between treatment groups or healthcare providers, in addition to being useful for risk prediction models. Based on the study’s results, we will suggest which approaches should be considered by other researchers with similar large datasets depending on the intended purpose of the model developed.

We expect that researchers who aim to develop a parsimonious prognostic index specifically for the outcome and population being studied could use the approach proposed under ‘First analysis’ above. This index—a single measure defined by a weighted number of conditions—could then be incorporated into future models to help reduce confounding or improve the performance of prediction models. Alternatively, the list of conditions that define the final index (without the weights) could be used as separate predictor variables in subsequent models. The index could therefore be used as the Charlson and Elixhauser indices have been; the difference is that the new approach produces an index that is optimal for a specific outcome and population.

Researchers who do not intend to develop a parsimonious index may prefer one of the approaches given under ‘Second analysis’. These approaches produce models that have a large number of diagnostic parameters and are more difficult to interpret but may predict outcomes better. A single measure can be produced as the predicted log-odds of death based on the diagnostic predictors in the model. This measure could then be included in future models as before. Alternatively, it may be preferable to estimate a model with diagnostic information and all other predictors simultaneously (using the same approaches), such as when it is acceptable for the final model to represent the conditions as a large number of separate terms.

Each of the approaches involves a large number of predictors such that large sample sizes are needed to develop the models. The adequate sample size depends on several factors including the frequencies of the outcome and co-existing conditions in the population. Guidance for determining adequate sample sizes is available elsewhere.^77^ Researchers with smaller datasets can still apply the models developed in larger studies to their own data, including those developed for acute myocardial infarction, hip fracture, and major colorectal cancer surgery in this study.

## Data Availability

No additional data are available.

## Contributors

TEC conceived the initial idea for the work and designed the study with substantial contributions from DAC, LDS, and JvdM. TEC will conduct the analysis for this study. TEC drafted the initial study protocol. All authors commented on draft manuscripts and approved this version of the protocol.

## Funding

This work was supported by a Medical Research Council fellowship awarded to TEC (grant number: MR/S020470/1).

## Competing interests

None to declare.

## Ethical approval

Hospital Episode Statistics and Civil Registrations Data were made available by NHS Digital; approval for the use of the data was obtained from NHS Digital as part of the standard approval process (digital.nhs.uk).

## Appendix 1. Tuning parameters of gradient boosted trees

We will use the caret^78^ package in R to tune the gradient boosted trees fitted by the XGBoost package. The final model will use the values of tuning parameters that minimised the negative log-likelihood for binary outcome variables. The parameters and the values tested will be:

| Parameter | Description | Values tested |
| --- | --- | --- |
| Number of iterations | Maximum number of boosting iterations | 1 and 25 to 500<br>(in steps of 25) |
| Learning rate | Scales the contribution of each tree's predictions by this value when added to the existing model (shrinkage) | 0.05 and 0.1 |
| Maximum tree depth | Highest level of predictor interactions allowed in a tree | 3 and 5 |
| Minimum node weight | Stops tree splitting if the sum of observation weights in a node is less than this parameter | 100 and 1000 |
| Subsample fraction | Fraction of observations in the training dataset randomly chosen to fit the next tree | 0.5 and 1 |

The parameter values to test were chosen partly based on the default values of the XGBoost package as well as the gbm package (which also implements gradient boosted trees). For each parameter, we will test values that are as or more conservative than the defaults in these packages to reduce the risk of overfitting. More conservative values correspond to fewer boosting iterations; lower learning rates, tree depths, and subsampling fractions; and greater minimum node weights. Overall, 336 unique combinations of the parameter values will be tested.

## Notes

### Competing Interest Statement

The authors have declared no competing interest.

## References

1. Charlson ME, Pompei P, Ales KL, MacKenzie CR. A new method of classifying prognostic comorbidity in longitudinal studies: development and validation. J Chronic Dis 1987;40(5):373–83.

2. Web of Science. Available from: www.webofknowledge.com.

3. Elixhauser A, Steiner C, Harris DR, Coffey RN. Comorbidity measures for use with administrative data. Medical Care 1998;36(1):8–27.

4. Zimmermann C, Swami N, Krzyzanowska M, Hannon B, Leighl N, Oza A, et al. Early palliative care for patients with advanced cancer: a cluster-randomised controlled trial. Lancet 2014;383(9930):1721–30.

5. Pro CI, Yealy DM, Kellum JA, Huang DT, Barnato AE, Weissfeld LA, et al. A randomized trial of protocol-based care for early septic shock. N Engl J Med 2014;370(18):1683–93.

6. Cohen HJ, Feussner JR, Weinberger M, Carnes M, Hamdy RC, Hsieh F, et al. A controlled trial of inpatient and outpatient geriatric evaluation and management. N Engl J Med 2002;346(12):905–12.

7. Schermerhorn ML, O’Malley AJ, Jhaveri A, Cotterill P, Pomposelli F, Landon BE. Endovascular vs. open repair of abdominal aortic aneurysms in the Medicare population. N Engl J Med 2008;358(5):464–74.

8. Baxter NN, Goldwasser MA, Paszat LF, Saskin R, Urbach DR, Rabeneck L. Association of colonoscopy and death from colorectal cancer. Ann Intern Med 2009;150(1):1–8.

9. Shahinian VB, Kuo YF, Freeman JL, Goodwin JS. Risk of fracture after androgen deprivation for prostate cancer. N Engl J Med 2005;352(2):154–64.

10. Birkmeyer JD, Siewers AE, Finlayson EV, Stukel TA, Lucas FL, Batista I, et al. Hospital volume and surgical mortality in the United States. N Engl J Med 2002;346(15):1128–37.

11. Werner RM, Bradlow ET. Relationship between Medicare’s hospital compare performance measures and mortality rates. JAMA 2006;296(22):2694–702.

12. Jha AK, Orav EJ, Li Z, Epstein AM. The inverse relationship between mortality rates and performance in the Hospital Quality Alliance measures. Health Aff (Millwood) 2007;26(4):1104–10.

13. Piccirillo JF, Tierney RM, Costas I, Grove L, Spitznagel EL, Jr. Prognostic importance of comorbidity in a hospital-based cancer registry. JAMA 2004;291(20):2441–7.

14. Yourman LC, Lee SJ, Schonberg MA, Widera EW, Smith AK. Prognostic indices for older adults: a systematic review. JAMA 2012;307(2):182–92.

15. Lee DS, Austin PC, Rouleau JL, Liu PP, Naimark D, Tu JV. Predicting mortality among patients hospitalized for heart failure: derivation and validation of a clinical model. JAMA 2003;290(19):2581–7.

16. Quan H, Sundararajan V, Halfon P, Fong A, Burnand B, Luthi JC, et al. Coding algorithms for defining comorbidities in ICD-9-CM and ICD-10 administrative data. Med Care 2005;43(11):1130–9.

17. xvan Walraven C, Austin PC, Jennings A, Quan H, Forster AJ. A Modification of the Elixhauser Comorbidity Measures into a Point System for Hospital Death Using Administrative Data. Medical Care 2009;47(6):626–33.

18. Sharabiani MT, Aylin P, Bottle A. Systematic review of comorbidity indices for administrative data. Med Care 2012;50(12):1109–18.

19. Gagne JJ, Glynn RJ, Avorn J, Levin R, Schneeweiss S. A combined comorbidity score predicted mortality in elderly patients better than existing scores. J Clin Epidemiol 2011;64(7):749–59.

20. Stanley J, Sarfati D. The new measuring multimorbidity index predicted mortality better than Charlson and Elixhauser indices among the general population. J Clin Epidemiol 2017;92:99–110.

21. Sarfati D, Gurney J, Stanley J, Salmond C, Crampton P, Dennett E, et al. Cancer-specific administrative data–based comorbidity indices provided valid alternative to Charlson and National Cancer Institute Indices. Journal of Clinical Epidemiology 2014;67(5):586–95.

22. Thombs BD, Singh VA, Halonen J, Diallo A, Milner SM. The effects of preexisting medical comorbidities on mortality and length of hospital stay in acute burn injury: evidence from a national sample of 31,338 adult patients. Ann Surg 2007;245(4):629–34.

23. Holman CD, Preen DB, Baynham NJ, Finn JC, Semmens JB. A multipurpose comorbidity scoring system performed better than the Charlson index. J Clin Epidemiol 2005;58(10):1006–14.

24. Einav L, Finkelstein A, Mullainathan S, Obermeyer Z. Predictive modeling of U.S. health care spending in late life. Science 2018;360(6396):1462–5.

25. Elfiky AA, Pany MJ, Parikh RB, Obermeyer Z. Development and Application of a Machine Learning Approach to Assess Short-term Mortality Risk Among Patients With Cancer Starting Chemotherapy. JAMA Netw Open 2018;1(3):e180926. doi: 10.1001/jamanetworkopen.2018.0926.

26. Rahimian F, Salimi-Khorshidi G, Payberah AH, Tran J, Ayala Solares R, Raimondi F, et al. Predicting the risk of emergency admission with machine learning: Development and validation using linked electronic health records. PLOS Medicine 2018;15(11):e1002695. doi: 10.1371/journal.pmed.1002695.

27. Avati A, Jung K, Harman S, Downing L, Ng A, Shah NH. Improving palliative care with deep learning. BMC Med Inform Decis Mak 2018;18(Suppl 4):122. doi: 10.1186/s12911-018-0677-8.

28. Jung K, Sudat SEK, Kwon N, Stewart WF, Shah NH. Predicting Need for Advanced Illness or Palliative Care In A Primary Care Population Using Electronic Health Record Data. J Biomed Inform 2019:103115. doi: 10.1016/j.jbi.2019.103115.

29. National Clinical Guideline Centre. The Management of Hip Fracture in Adults. 2011. Available from: https://www.nice.org.uk/guidance/cg124/evidence/full-guideline-pdf-183081997.

30. Chatterton BD, Moores TS, Ahmad S, Cattell A, Roberts PJ. Cause of death and factors associated with early in-hospital mortality after hip fracture. Bone Joint J 2015;97-B(2):246–51.

31. Metcalfe D, Zogg CK, Judge A, Perry DC, Gabbe B, Willett K, et al. Pay for performance and hip fracture outcomes: an interrupted time series and difference-in-differences analysis in England and Scotland. Bone Joint J 2019;101-B(8):1015–23.

32. Asaria P, Elliott P, Douglass M, Obermeyer Z, Soljak M, Majeed A, et al. Acute myocardial infarction hospital admissions and deaths in England: a national follow-back and follow-forward record-linkage study. The Lancet Public Health 2017;2(4):e191–e201. doi: 10.1016/s2468-2667(17)30032-4.

33. Bottle A, Aylin P. Comorbidity scores for administrative data benefited from adaptation to local coding and diagnostic practices. J Clin Epidemiol 2011;64(12):1426–33.

34. Nuffield Trust. Stroke and heart attack mortality. Available from: https://www.nuffieldtrust.org.uk/resource/stroke-and-heart-attack-mortality.

35. Dobbins TA, Creighton N, Currow DC, Young JM. Look back for the Charlson Index did not improve risk adjustment of cancer surgical outcomes. J Clin Epidemiol 2015;68(4):379–86.

36. Boyle J, Braun M, Hill J, Kuryba A, van der Meulen J, Walker K, et al. National Bowel Cancer Audit Annual Report 2018. Available from: https://www.nboca.org.uk/content/uploads/2018/12/NBOCA-annual-report2018.pdf.

37. Herbert A, Wijlaars L, Zylbersztejn A, Cromwell D, Hardelid P. Data Resource Profile: Hospital Episode Statistics Admitted Patient Care (HES APC). Int J Epidemiol 2017;46(4):1093–i. doi: 10.1093/ije/dyx015.

38. Metcalfe A, Neudam A, Forde S, Liu M, Drosler S, Quan H, et al. Case definitions for acute myocardial infarction in administrative databases and their impact on in-hospital mortality rates. Health Serv Res 2013;48(1):290–318.

39. McCormick N, Lacaille D, Bhole V, Avina-Zubieta JA. Validity of myocardial infarction diagnoses in administrative databases: a systematic review. PLoS One 2014;9(3):e92286. doi: 10.1371/journal.pone.0092286.

40. Toson B, Harvey LA, Close JC. The ICD-10 Charlson Comorbidity Index predicted mortality but not resource utilization following hip fracture. J Clin Epidemiol 2015;68(1):44–51.

41. Royal College of Physicians. National Hip Fracture Database (NHFD) annual report 2016. 2016. Available from: https://www.nhfd.co.uk/report2016.

42. Burns EM, Bottle A, Aylin P, Darzi A, Nicholls RJ, Faiz O. Variation in reoperation after colorectal surgery in England as an indicator of surgical performance: retrospective analysis of Hospital Episode Statistics. BMJ 2011;343:d4836. doi: 10.1136/bmj.d4836.

43. Byrne BE, Mamidanna R, Vincent CA, Faiz O. Population-based cohort study comparing 30- and 90-day institutional mortality rates after colorectal surgery. Br J Surg 2013;100(13):1810–7.

44. Morris EJ, Taylor EF, Thomas JD, Quirke P, Finan PJ, Coleman MP, et al. Thirty-day postoperative mortality after colorectal cancer surgery in England. Gut 2011;60(6):806–13.

45. Redaniel MT, Martin RM, Blazeby JM, Wade J, Jeffreys M. The association of time between diagnosis and major resection with poorer colorectal cancer survival: a retrospective cohort study. BMC Cancer 2014;14(1):642.

46. World Health Organization. International Statistical Classification of Diseases and Related Health Problems - 10th revision (5th edition). 2016. Available from: https://icd.who.int/browse10/Content/statichtml/ICD10Volume2_en_2016.pdf.

47. Yurkovich M, Avina-Zubieta JA, Thomas J, Gorenchtein M, Lacaille D. A systematic review identifies valid comorbidity indices derived from administrative health data. J Clin Epidemiol 2015;68(1):3–14.

48. Office for National Statistics. Deaths. Available from: https://www.ons.gov.uk/peoplepopulationandcommunity/birthsdeathsandmarriages/deaths.

49. Krumholz HM, Coppi AC, Warner F, Triche EW, Li SX, Mahajan S, et al. Comparative Effectiveness of New Approaches to Improve Mortality Risk Models From Medicare Claims Data. JAMA Netw Open 2019;2(7):e197314. doi: 10.1001/jamanetworkopen.2019.7314.

50. Ministry of Housing, Communities & Local Government,. English indices of deprivation. Available from: https://www.gov.uk/government/collections/english-indices-of-deprivation.

51. Harrell FE, Jr. Regression Modeling Strategies. New York: Springer; 2015.

52. Ambler G, Brady AR, Royston P. Simplifying a prognostic model: a simulation study based on clinical data. Stat Med 2002;21(24):3803–22.

53. Lawless JF, Singhal K. Efficient Screening of Nonnormal Regression-Models. Biometrics 1978;34(2):318–27.

54. Simard M, Sirois C, Candas B. Validation of the Combined Comorbidity Index of Charlson and Elixhauser to Predict 30-Day Mortality Across ICD-9 and ICD-10. Med Care 2018;56(5):441–7.

55. Hastie T, Tibshirani R. Generalized Additive Models. London: Chapman & Hall; 1990.

56. Hastie TJ. Generalized additive models. In: Chambers JM, Hastie TJ, editors. Statistical Models in S. London: Chapman & Hall; 1993.

57. Friedman JH. Greedy function approximation: A gradient boosting machine. Annals of Statistics 2001;29(5):1189–232.

58. Friedman J, Hastie T, Tibshirani R. Additive logistic regression: a statistical view of boosting (With discussion and a rejoinder by the authors). The Annals of Statistics 2000;28(2):337–407.

59. Friedman JH. Stochastic gradient boosting. Computational Statistics & Data Analysis 2002;38(4):367–78.

60. Chen T, Guestrin C. XGBoost: A Scalable Tree Boosting System. arXiv 2016. doi: 10.1145/2939672.2939785.

61. Kuhn M, Johnson K. Applied Predictive Modeling. New York: Springer; 2013.

62. James G, Witten D, Hastie T, Tibshirani R. An Introduction to Statistical Learning with Applications in R. New York: Springer; 2013.

63. Chen T, He T, Benesty M. XGBoost R Tutorial. Available from: https://cran.r-project.org/web/packages/xgboost/vignettes/xgboostPresentation.html.

64. Brier GW. Verification of Forecasts Expressed in Terms of Probability. Monthly Weather Review 1950;78(1):1–3.

65. Steyerberg EW, Vickers AJ, Cook NR, Gerds T, Gonen M, Obuchowski N, et al. Assessing the performance of prediction models: a framework for traditional and novel measures. Epidemiology 2010;21(1):128–38.

66. Harrell FE, Jr., Califf RM, Pryor DB, Lee KL, Rosati RA. Evaluating the yield of medical tests. JAMA 1982;247(18):2543–6.

67. Cox DR. Two further applications of a model for binary regression. Biometrika 1958;45(3-4):562–5.

68. Austin PC, Steyerberg EW. The Integrated Calibration Index (ICI) and related metrics for quantifying the calibration of logistic regression models. Stat Med 2019;0(0). doi: 10.1002/sim.8281.

69. Harrell FE, Jr., Lee KL, Mark DB. Multivariable prognostic models: issues in developing models, evaluating assumptions and adequacy, and measuring and reducing errors. Stat Med 1996;15(4):361–87.

70. Moons KG, Altman DG, Reitsma JB, Ioannidis JP, Macaskill P, Steyerberg EW, et al. Transparent Reporting of a multivariable prediction model for Individual Prognosis or Diagnosis (TRIPOD): explanation and elaboration. Ann Intern Med 2015;162(1):W1–73. doi: 10.7326/M14-0698.

71. Steyerberg EW. Clinical Prediction Models. New York: Springer; 2009.

72. Healthcare Cost and Utilization Project. Clinical Classifications Software (CCS) for ICD-9-CM. Available from: https://www.hcup-us.ahrq.gov/toolssoftware/ccs/ccs.jsp#download.

73. NHS Digital. Summary Hospital-level Mortality Indicator (SHMI): ICD-10 to SHMI diagnosis group lookup table. Available from: https://digital.nhs.uk/data-and-information/publications/ci-hub/summary-hospital-level-mortality-indicator-shmi.

74. Verweij PJ, Van Houwelingen HC. Penalized likelihood in Cox regression. Stat Med 1994;13(23-24):2427–36.

75. StataCorp. Stata Statistical Software: Release 15. College Station, TX: StataCorp LLC; 2017.

76. R Core Team. R: A language and environment for statistical computing. Vienna, Austria: R Foundation for Statistical Computing; 2017. Available from: https://www.r-project.org/.

77. Riley RD, Snell KI, Ensor J, Burke DL, Harrell FE, Jr., Moons KG, et al. Minimum sample size for developing a multivariable prediction model: PART II - binary and time-to-event outcomes. Stat Med 2018. doi: 10.1002/sim.7992.

78. Kuhn M. Building Predictive Models in R Using the caret Package. Journal of Statistical Software 2008;28(5):1–26.

